# Personalized Feeding Infusion Rate of Nutrition to Improve Glycemic Variability Control in Critical Care Patients

**DOI:** 10.1101/2021.12.23.21268339

**Authors:** Rammah Abohtyra

## Abstract

Control-based algorithms in the intensive care unit (ICU) patients have been developed to deliver a sufficient amount of insulin, but optimizing the rate of feeding of nutrition in ICU patients to improve glycemic variability control has not been done yet. Continuous feeding is commonly used for nutrition in critically ill patients who cannot be fed orally to maintain a normal blood sugar concentration, but optimizing its rate, for these individuals, is needed to avoid the adverse outcomes caused by medications such as insulin. This paper develops a control-based algorithm combines a predictive control algorithm with a revised nonlinear compartmental model used in the ICU to design personalized feeding function rates to improve patient glycemic variability. Our control algorithm is robust and acts very quickly to avoid medical intervention effects.

## I. INTRODUCTION

Patients in the Intensive Care Unit (ICU), including non-diabetic patients, have high glycemic variability including Hyperglycemia (high blood sugar *>* 140 mg/dl) and Hypoglycemia (low blood sugar *<* 40 mg/dl), which in turn are associated with increased morbidity and mortality [1], [2], [3], [4]. A recent report indicated that Hyperglycemia occurs in 22%-46% of non-diabetic ICU patients [5]. Hypoglycemia which occurs in 10.1% of ICU patients has a more significant concern in ICU patients [6], [2]. Delivering an inadequate amount of insulin is the main cause of hypoglycemia [7].

Continuous feeding (enteral nutrition), which consists of nutritional infusion used at a constant rate, is selected to be the standard method of nutrition for patients in ICU [8], [9], with several benefits, including improves gut function, decreases infections, and lower mortality [10], but optimizing this method has not been done yet. For medical reasons, the administrated continuous feeding in the ICU should be initiated at a rate of 10-20 ml/h and then gradually increased to the target rate [11]. In ICU patients with acute kidney injury, it has been recommended that to reduce kidney failure morbidity, those individuals should be placed under continuous feeding to receive an amount of protein-restricted by body weight (1.2-2 g/kg body weight per day) [12].

Under continuous feeding, ultradian oscillations of plasma glucose, glycemic variability, with periods of 50-200 min, initially discovered by Hansen in [13], have been shown in non-diabetic patients [14], [15]. Damped, slow, and unregulated glucose oscillations have been also detected in patients with diabetes. [16].

Multiple models of plasma glucose regulation, with different levels of and fidelity of physiological representation, demonstrate ultradian oscillations under constant feeding [17], [18], [19]. The underlying mechanisms of these oscillations are based on different hypotheses.

Control-based algorithms with a closed-loop action, including Model Predictive Control (MPC), have been developed to regulate blood glucose dynamics in ICU settings [20], [21], [22]. All these closed-loop algorithms have been developed to deliver insulin without nurse input. This paper however develops an MPC-based algorithm to optimize the nutrition feeding rates for ICU patients and accounts for body size and unknown input medications such as insulin, the leading cause of hypoglycemia.

The paper is organized as follows. In Section II, a revised nonlinear model is introduced. The proposed control algorithm is described in Section III. Our simulation results are demonstrated in Section IV. A brief discussion is provided in Section V, and Conclusions and future plans are included in Section VI.

## II. Revised ICU Model Description

The Ultradian model developed by Sturis et al. [17] is considered in this work. This nonlinear multi-compartmental model oscillates under constant feeding due to a delayed insulin signaling cascade that triggers the liver to produce hepatic glucose. The original model (see [17], [23]) uses state variables, the total amount of insulin (*I*_*p*_ mU) and total amount of glucose (*G*_*p*_ mg) in the plasma compartment, and the total amount of insulin in the peripheral (interstitial) compartment (*I*_*i*_ mU); and the delayed insulin signaling to the liver are given by three cascade of insulin variables (total amount) *H*_1_, *H*_2_, and *H*_3_ (mU).

Since in the clinical practices measurements are taken in concentrations, we express the model in terms of compartment-wise concentrations of insulin and glucose as these are the relevant and clinically measured physiological variables. Additionally, in order to introduce a patient-specific parameter, we explicitly use the patient size Ω_*m*_ (non-dimensional), to express the peripheral compartment size 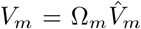 where 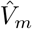, is the nominal peripheral (muscle) size in the original model [17]. For this work all other parameters match those in [17]. The recasted equations of the model are given by

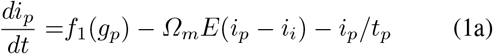

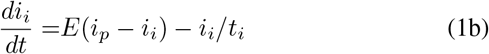

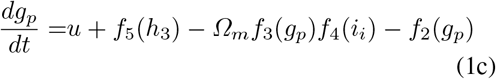

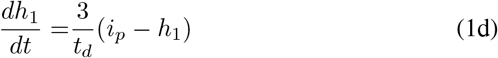

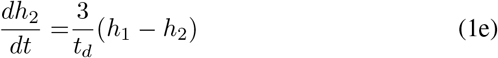

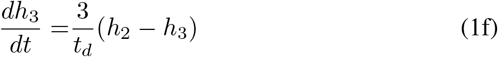

where *i*_*p*_ = *I*_*p*_*/V*_*p*_, *g*_*p*_ = *G*_*p*_*/V*_*g*_, *i*_*i*_ = *I*_*i*_*/V*_*m*_, are the concentrations of the plasma insulin, plasma glucose, and interstitial insulin respectively; *h*_1_, *h*_2_, and *h*_3_ describe the delayed insulin signaling variables in concentrations, which are computed as *h*_1_ = *H*_1_*/V*_*p*_, *h*_2_ = *H*_2_*/V*_*p*_, and *h*_3_ = *H*_3_*/V*_*p*_; *u* is an exogenous glucose infusion rate to be controlled; and where *V*_*p*_, (distribution volume for plasma insulin) *V*_*g*_ (distribution volume for plasma glucose), and *V*_*m*_ (interstitial space) are the compartmental sizes for this model; *f*_1_ defines the insulin production rate, *f*_3_*f*_4_ define the muscle absorption rate, *f*_2_ defines the brain absorption rate, and *f*_5_ defines the hepatic production rate. The mathematical equations of these production rates are given in Table I. The nominal parameters of the model are depicted in Table II.

**TABLE I.**
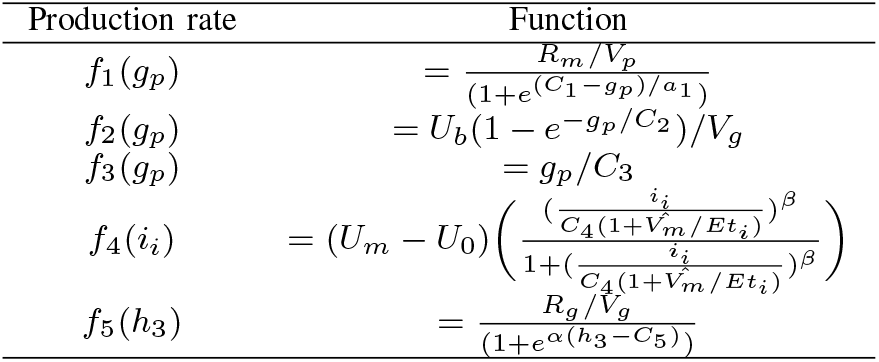
The model production rates are given by these functions.

**TABLE II.** The nominal parameters of the ultradian model, as reported in [23].

| parameter | value | parameter | value |
| --- | --- | --- | --- |
| $V_p$ (L) | 3 | $U_b$ (mg/min) | 72 |
| $\hat{V}_m$ (L) | 11 | $C_2$ (mg/L) | 144 |
| $V_g$ (L) | 10 | $C_3$ (mg/L) | 1000 |
| $E_0$ (L/min) | 0.2 | $U_0$ (mg/min) | 40 |
| $t_p$ (min) | 6 | $U_m$ (mg/min) | 940 |
| $t_i$ (min) | 100 | $\beta$ | 1.77 |
| $t_d$ (min) | 36 | $C_4$ (mg/L) | 80 |
| $R_m$ (mU/min) | 210 | $R_g$ (mU/min) | 180 |
| $a_1$ (mg/L) | 300 | $\alpha$ (L/mU) | 0.29 |
| $C_1$ (mg/L) | 2000 | $C_5$ (mU/L) | 26 |

Note that because the diffusive constant *E*, which appears in (1a) and (1b) is proportional to the compartmental muscle surface, we scale it by the relative muscle size Ω_*m*_; however, using 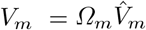, the term Ω_*m*_ is canceled out in (1b).

In Fig. 1, we illustrate the effect of the muscle size (Ω_*m*_: 0.6-1.6) on the glucose dynamics (oscillation amplitude and feeding rate). We note that increase the muscle size will require a higher feeding rate to keep the same blood glucose level, and yields changes in amplitude of oscillation and oscillation stability. The next section demonstrates an optimal design for the feeding rate as the body size changes.

**Fig. 1.**
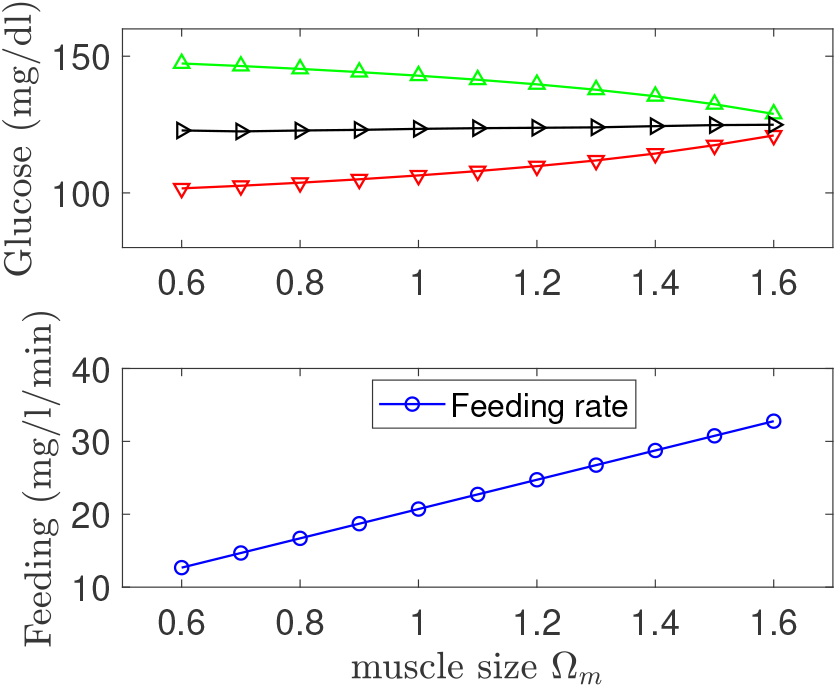
Top panel: Average glucose oscillation computed at a steady-state glucose level (125 mg/dl), represented by average upper peaks (green), average mean value (black), and average lower peaks (red) (mg/dl), versus different muscle sizes. Bottom panel: steady-state feeding rates (mg/l/min) computed at different muscle sizes.

## III. Model-Predictive Control Algorithm for Feeding Rate Control

In Fig. 2, we illustrate a schematic diagram for the proposed closed-loop Model-Predictive Control (MPC) algorithm. The components of this algorithm will be described in detail in this section.

**Fig. 2.**
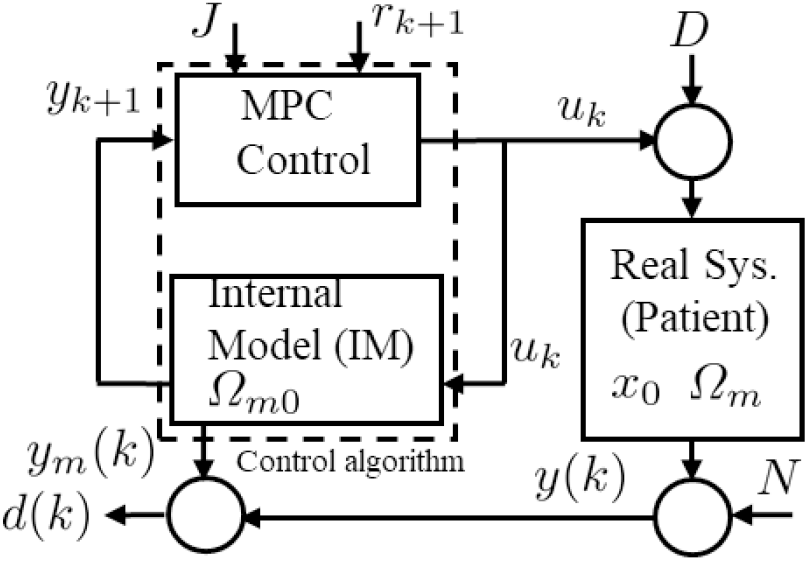
Closed-loop MPC control algorithm with a state observer scheme (Kalman filter).

The model-based Predictive Control improves glycemic control in the ICU by providing the service of counting the effect of current and future control inputs (i.e., delivered carbohydrate and insulin rates) on the future outputs (i.e., blood glucose) by imposing clinical constraints on the inputs and outputs of the system. It requires solving an optimal control problem, with chosen input and output horizons, at each time instant *T*, after which only the first control value (i.e., carbohydrate infusion or insulin rate for the next time instant) of the optimal input sequence is applied to the actual system (i.e., patient).

In the ICU, the MPC application has been illustrated primarily to optimize the insulin injection [20], [22].

In this work, the delivered glucose infusion flow is the desired input to be controlled, while insulin injection and glucose boluses are considered to be unknown medical interventions.

We refer to the “real system,” in Fig 2, by real patients, with arbitrary muscle sizes, and these muscle sizes are unknown to the controller.

In the rest of the paper, we assume that the nonlinear mathematical model (1a)-(1f), with nominal size *Ω*_*m*_ ≡ 1, is the best model of the patient dynamics. We linearize this nonlinear model (*Ω*_*m*_ = 1) to generate the internal model (IM), in Fig 2, around a specific fixed point solution 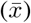 of the nonlinear model. The fixed point solution is chosen based on the control objective, which means if the control law is to stabilize the plasma glucose oscillation at 100 mg/dl, then we approximate the nonlinear model around 100 mg/dl to create IM.

We use the state-space (around this point) to estimate IM’s parameters, but we skip this step as a future plan of this paper.

At a sampling time *T*_*s*_ (min), the discrete-time version of IM is given by these model equations

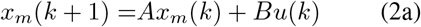

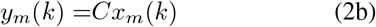

where *A, B, C* are the model matrices which are known.

In our control algorithm, the IM is simulated, using the feeding input *u*, in parallel with the real system “patient” (nonlinear model with unknown muscle size Ω_*m*_ to the controller) to produce a simulated blood glucose *y*_*m*_(*k*), while, the nonlinear model (with unknown muscle size) generates “real” glucose measurements (i.e., patient measurements), which take place each *Ts* (e.g., 5-15min), denoted by *y*(*k*). We use *y*_*m*_(*k*) to predict the offset between the real measurements and IM through *d*(*k*) = *y*(*k*) − *y*_*m*_(*k*), which captures the actual unknown disturbance (medications) and parameter uncertainty. The mismatch between the real measurements and IM is significantly reduced using the Kalman filter by updating the states of IM with the current measurement information from the patient. Therefore, accurate predictions using the updated model will be obtained [24]. Due to a lack of space, the prediction equations of IM, *y*_*m*_(*k* + 1), using the discrete-time model (2), can be found elsewhere [25].

The predictions of the real system (patient), *y*(*k* + 1), are formulated using the predictions of IM and the prediction of the offset *d*(*k*) through *y*(*k* + 1) = *y*_*m*_(*k* + 1) + *d*(*k*), assuming that *d*(*k* + 1) = *d*(*k*). These predictions will be incorporated within the MPC controller to solve an open-loop optimization problem online using a quadratic objective function to generate future feeding input rates *u*(*k* +1). As a close-loop action, the first value of *u* is applied in the algorithm at each sampling time.

The objective function that needs to be minimized is described as follows:

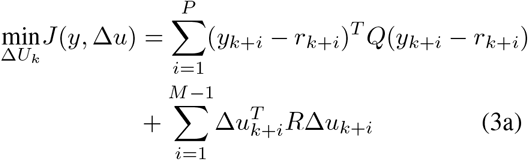

Subject to

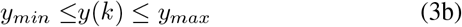

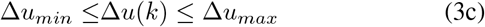

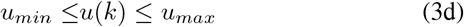

where Δ*U*_*k*_ = [Δ*u*_*k*_ *…* Δ*u*_*k*+*M*−1_], in which Δ*u*_*k*+1_ = *u*_*k*+1_ − *u*_*k*−1_, denotes a vector containing all changes in the future feeding rates, within a given input horizon *M*. This objective function comprises a trade-off between allowable changes in input rates and deviations from the desired glycemic level. The design parameters of the MPC are the weighting matrices *Q* and *R*, the control horizon (*M*), and the prediction horizon *P*.

The upper and lower constraints of glucose, *y*_*min*_ and *y*_*max*_ in (3b), keep the average blood glucose level of the nonlinear model within this range [40-140] mg/dl, by optimizing the objective function (3a) for a given target blood glucose trajectory.

The constraints *u*_*min*_ and *u*_*max*_ in (3d) limit the feeding rate *u* based on Δ*u*, which itself has given clinical bounds (Δ*u*_*min*_, Δ*u*_*max*_) imposed by the second constraint. These bounds will guarantee that the feeding pump does not go too high or too low. All the above linear inequalities (constraints) are transformed in a matrix form and incorporated in the objective function (3a).

## IV. Simulation Results

In this section, the proposed MPC algorithm will be implemented and tested in the presence of measurement noise (white noise with standard deviation *σ* = 5 mg/dl) to improve glycemic variability, avoid a simulated insulin intervention, and track control objectives (glucose trajectories). The minimum glucose level implemented in the MPC algorithm was *y*_*min*_ = 60 mg/dl, which is a slightly above the hypoglycemia level (40 mg/dl), and the maximum glucose values was *y*_*max*_ = 140 mg/dl. The maximum constraints of the feeding rate were selected as Δ*u*_*min*_ = 0, Δ*u*_*max*_ = 1 mg/dl/min, while, the limits of the feeding input were *u*_*min*_ = 0, *u*_*max*_ = 2.5 mg/dl/min [18].

The IM was constructed by approximating the nonlinear model around the control objective (glucose level) 125 mg/dl, with the nominal muscle size (Ω_*m*0_ = 1).

The parameters of the MPC algorithem were selected to achieve a good control performance. The lengths of prediction and input horizons were chosen to be *P* = 15 and *M* = 5, while, the weighting matrices were *R* = 5, *Q* = 1.

To show the effect of the MPC control on glycemic variability around the plasma glucose 125 mg/dl, we simulate the nonlinear model, with nominal muscle size, using an average feeding function of nutrition at 2.5 mg/dl/min, to produce oscillatory glycemic variability behavior as shown in Fig. 3. The MPC was turned on at 500 min to stabilize the plasma glucose around 125 mg/dl and tracks 110 mg/dl (second control objective) and 125 mg/dl.

**Fig. 3.**
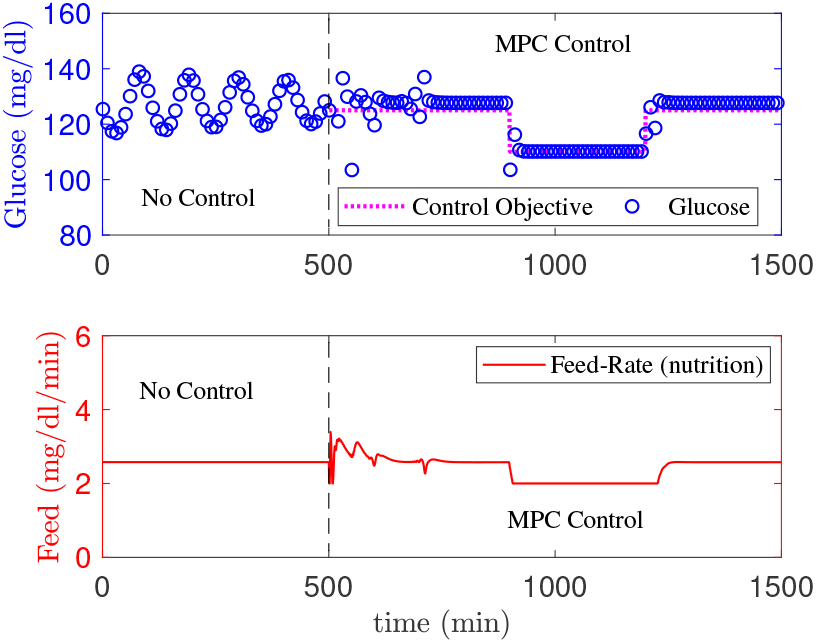
Top panel: a comparison between glycemic variability (oscillation) before and after control MPC is shown. The oscillation was stabilized at 125 mg/dl (objective control) when the MPC was turned on at 500 min. At 800 min, the control objective was changed to track 110 mg/dl. Bottom panel: feeding rate of nutrition before and during the control.

The administration of medication (insulin injection) can also shift blood glucose levels. To test the performance of our control algorithm by adding an insulin infusion (unknown to the MPC) to the real system at 1000 min which illustrated in Fig 4. The injected insulin shifted the amplitude of the plasma glucose. As a consequence, the MPC responds quickly by increasing the feeding rates to manigate the shift in the glucose amplitude, and bring it back to the same amplitude level.

**Fig. 4.**
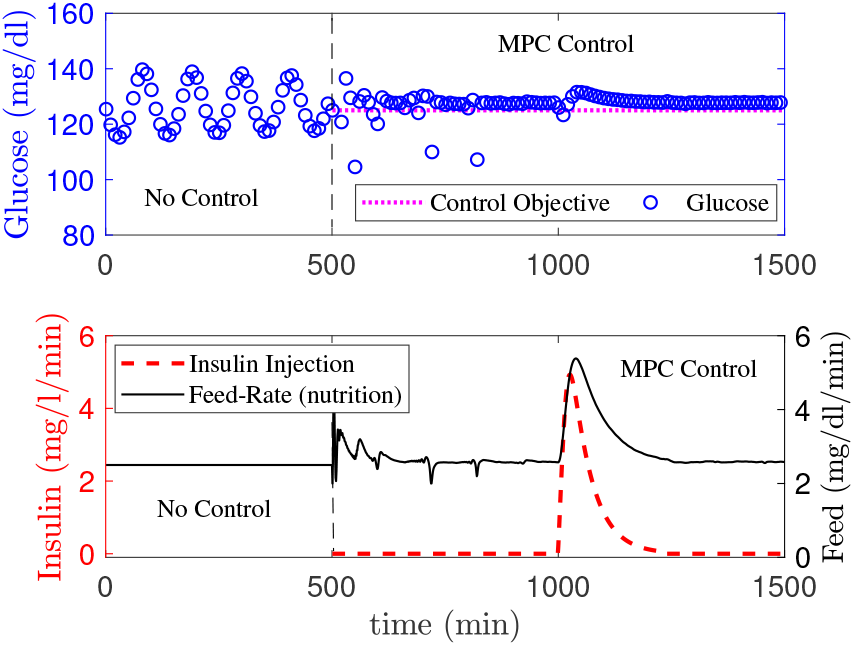
Top panel: performance of the MPC control algorithm to maintain plasma glucose at a target level (control objective) against unknown insulin intervention to the real system (patient), applied at 1000 min; Bottom panel, feeding rate was adjusted by the controller to overcome the reduction in the plasma glucose by the insulin injection.

To generate personalized feeding rates when the muscle size changes, the MPC control algorithm was simulated with the real system (patient) (IM computed at a fixed-point solution, 105 mg/dl) with different muscle sizes (muscle size increased by 25% and 75%), to track a target glucose trajectory (105 mg/dl, 95 mg/dl, 105 mg/dl), as shown in Fig 5 and 6, upper panels.

**Fig. 5.**
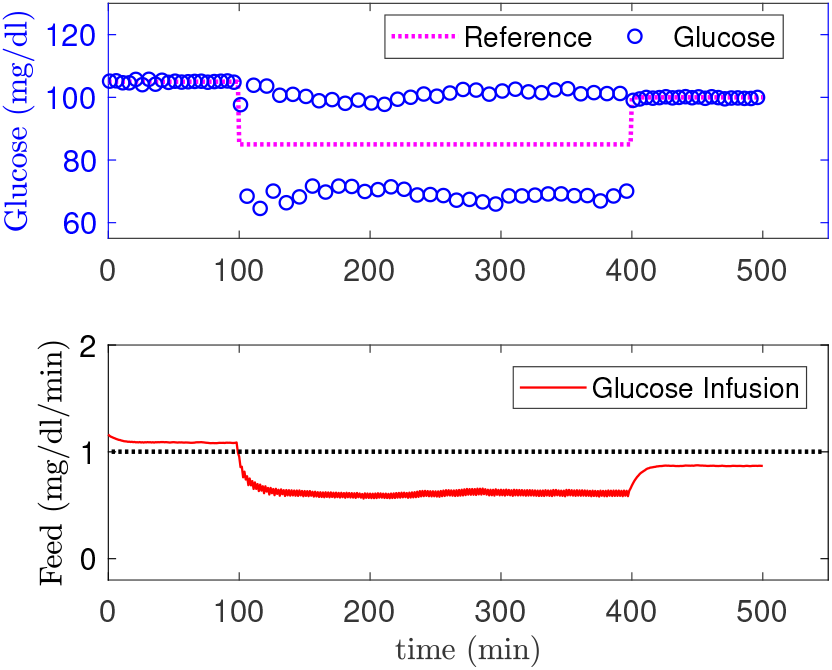
Glucose variability is stabilized at 105 mg/dl (dots magenta), using the MPC with IM computed at 105 mg/dl, while, the oscillation is kept within between 70-105 mg/dl for 95 mg/dl control objective. Lower panel: the proposed feeding function from the MPC algorithm (red), with parameters: muscle sized increased by 25% (Ω_*m*_ = 1.25), *R* = 5.

**Fig. 6.**
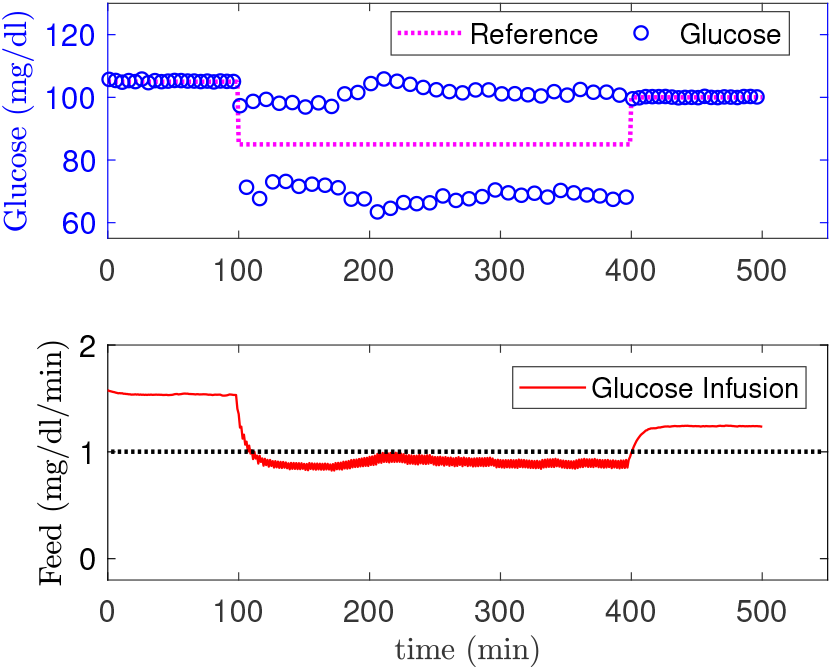
Glucose variability (with muscle sized 75% increased, Ω_*m*_ = 1.75) is stabilized at 105 mg/dl (dots magenta), using the MPC with IM computed at 105 mg/dl, while, the oscillation is kept within 70-105 mg/dl for 95 mg/dl control objective. Lower panel: the proposed feeding function for the MPC algorithm (red),, *R* = 10.

To provide a basis for comparison between the feeding rates proposed by the MPC when the muscle size was increased, 25% (Ω_*m*_ = 1.25) and 75% (Ω_*m*_ = 1.75) respectively, we draw a horizontal line at the rate 1 mg/dl/min as shown in Fig 5 and 6, lower panels. The MPC proposed different feeding rates based on the muscle size to achieve the same control objective.

## V. Discussion

The ultradian model was recasted to track the concentrations across the compartments, and a relative body size was implemented to account for patient’s body size variability.

The MPC stabilized the plasma glucose at 105 mg/dl for the first control objective (105 mg/dl). Simultaneously, the MPC kept glycemic variability in both cases (Ω = 1.25 and Ω = 1.25) within the same variability range (70-105 mg/dl) without stabilizing the oscillation to 95 mg/dl because IM was made at 105 mg/dl. To stabilize the plasma glucose at 95 mg/dl, the IM should be built using 95 mg/dl, as a fixed point solution of the nonlinear model.

The proposed model-based predictive control with state estimation, using Kalman filter, has several advantages over standard MPC. The patient’s information provided to the MPC controller yields tighter control and adjust closed-loop performance. Utilizing the internal model has several benefits that the controller can estimate the states of the nonlinear model, formulate the predictions, and solve a quadratic optimization problem online.

The MPC control algorithm was designed to produce optimal feeding infusion rates that keep the glucose oscillations within the desired range (40-140 mg/dl) while avoiding medical interventions as unknown inputs to this model. The produced feeding rates are personalized rates based on the body size.

Although the MPC overcame the unknown medication disturbances, glucose, and insulin injections, the insulin’s effect on the oscillations was more significant and stayed for a longer time.

## VI. Conclusions and Future Works

### A. Conclusions

A successful model-based predictive control algorithm was developed for glycemic control. Our control algorithm acts quickly and improves glycemic variability at a desired plasma glucose level. This MPC algorithm was designed to produce personalized optimal feeding infusion rates based on patient’s body size, while avoiding unknown medical interventions. The algorithm’s performance proves its robustness as an essential feature for possible use in a real-life ICU setting.

### B. Future Works

Future work is directed to implement time-varying internal models to achieve multiple control objectives, a parameter estimate approach (such as a moving horizon estimator (MHE)), and incorporate more mechanistic glucose models, thus leading to further improvement in the closed-loop control algorithm’s performance.

## Data Availability

In this study there is No data, only computational work.

## VII. ACKNOWLEDGMENTS

This work is supported by grants from the National Institutes of Health (t (R01LM0127).

